# The FDA’s Proposed Rule on Laboratory-Developed Tests: Impacts on Clinical Laboratory Testing and Patient Care

**DOI:** 10.1101/2024.02.28.24303459

**Authors:** Leslie Smith, Lisa A. Carricaburu, Jonathan R. Genzen

## Abstract

In October 2023, the U.S. Food and Drug Administration (FDA) released a proposed rule to regulate laboratory-developed tests (LDTs) as medical devices. While approximately 6,700 public comments were submitted during the open comment period, there is not a reliable mechanism to quantify how clinical laboratorians as a sector perceive the proposed rule. To solicit quantifiable feedback on the FDA’s proposed rule, a ten-item questionnaire was developed and submitted to clinical laboratory customers of ARUP Laboratories, a national nonprofit clinical laboratory of the University of Utah Department of Pathology.

Of 503 clinical laboratory respondents, only 41 (8.2%) support the FDA’s proposed rule. 66.9% of respondents work in laboratories that perform LDTs and were therefore asked additional questions regarding the proposed rule. 83.9% of these respondents believe that the proposed rule will negatively impact their laboratories, while only 3.0% believe that they have the financial resources to pay for FDA user fees. 60.9% of respondents anticipate removing tests from their laboratory menus if the proposed rule is enacted, while an additional 33.2% indicated that they do not yet know. Only 11.2% of respondents believe that they would pursue FDA submissions for all of their existing LDTs if the final rule is enacted.

The vast majority of respondents (>80%) were either ‘extremely concerned’ or ‘very concerned’ about the impact of the proposed rule on patient access to essential testing, financial and personnel resources to comply, innovation, the FDA’s ability to implement the proposed rule, and send-out costs and test prices. Respondents indicated that they would rely heavily on reference laboratory partners for advocacy against the proposed rule, testing options, education, and consultation if the rule was enacted. Thematic analysis of open comments revealed strong opposition to the proposed rule and significant concern regarding negative impacts to patient care across clinical laboratory settings.

## Introduction

On October 3, 2023, the U.S. Food and Drug Administration (FDA) released a proposed rule that would regulate laboratory-developed tests (LDTs) as medical devices if enacted [1]. The FDA has defined LDTs as in vitro diagnostics (IVDs) that are “intended for clinical use and designed, manufactured and used within a single laboratory” [2]. While there is no legislative definition from Congress, nor any current federal regulations that create a legal definition of LDTs in the U.S., the FDA’s definition has been promulgated extensively in non-binding draft guidance documents and public-facing statements [3, 4].

LDTs are not mentioned in the Medical Device Amendments of 1976 [5] – the law that established the current framework for medical device regulation in the U.S. – nor were LDTs discussed in Congressional hearings prior to the law’s passage [6, 7]. Thus, there is significant legal uncertainty as to whether the FDA has the authority from Congress to advance the current proposed rule [8]. Regardless, FDA leadership has signaled a commitment to move forward with LDT oversight [9], and an April 2024 tentative date for finalization of the proposed rule was posted in the fall 2023 Unified Agenda from the Biden Administration [10, 11].

In the U.S., rulemaking from federal agencies must follow requirements outlined in the Administrative Procedure Act [12]. The FDA’s proposed rule on LDTs is considered ‘notice-and-comment’ rulemaking, where the public is notified of a proposed rule and provided with the opportunity to provide open comments, and the agency is required to consider this feedback from the public in its deliberation and rulemaking [13]. The public comment period for the FDA’s proposed rule was limited to 60 days, and it officially closed on December 4, 2023, with approximately 6,700 comments submitted [14]. The FDA declined numerous requests for extensions of the public comment period [15], however. These requests included a letter signed by leaders of 89 laboratories and professional organizations [16], as well as a request from the American Medical Association House of Delegates [17]. Extension of public comment periods can provide members of the public more time to evaluate the implication of a proposed rule, and they are often granted by agencies for rules with significant potential impact to the public. The FDA has hosted no public workshops or hearings on LDT oversight since 2015 [18].

An important aspect of the public comment process is to assist a federal agency in understanding whether different sectors of the public support or oppose a proposed rule and for which reasons. While the Federal Register’s comment submission system for the proposed rule on LDTs included a drop-down menu for submitters to select the industry, corporate type, career, or setting most applicable to them – presumably to assist federal agencies in categorizing comments by sector – ‘clinical laboratory’ or ‘clinical laboratorian’ were not available options, even though this setting and career type is most impacted by the proposed rule. As such, it will be difficult for the FDA to conduct an accurate quantitative analysis of public opinion in a key sector directly impacted by its proposed rule.

To provide a quantitative assessment of clinical laboratorian opinions on the FDA’s proposed rule and its anticipated impact on clinical laboratory operations and patient care, a survey was distributed in February 2024 to customer contacts of ARUP Laboratories. ARUP is a national, nonprofit clinical laboratory enterprise of the University of Utah Department of Pathology, with community hospital and academic medical center laboratory customers across all 50 states [19]. As such, recipients of this survey are directly involved in clinical laboratory testing and have occupational roles for which FDA-cleared/approved IVDs and LDTs are relevant and familiar. Survey results demonstrate that there is strong opposition among clinical laboratory respondents to the proposed rule and significant concerns regarding its anticipated negative impacts on clinical laboratory testing and patient care across laboratories.

## Materials and methods

A ten-item questionnaire regarding the potential impact of the FDA’s proposed rule on LDTs was developed (see Survey Questionnaire). The study received an exemption determination from the University of Utah Institutional Review Board (IRB 00174067). An invitation to participate in this survey was distributed to customer contact email addresses from ARUP’s customer relationship management system (CRM) (Salesforce; San Francisco, CA). Individual survey links could only be used for one submission. As ARUP Laboratories operates the University of Utah Health clinical laboratories and also serves as the health system’s primary reference laboratory, surveys were not distributed to either ARUP or University of Utah Health email addresses to avoid the potential for respondent bias regarding expectations of reference laboratory services. Contacts at national reference laboratories were also excluded to avoid similar bias regarding reference laboratory services. Research trial contacts were also excluded as not all studies require CLIA-certification. The survey was administered using the Experience Management XM platform (Qualtrics; Provo, UT), and no identifiers were collected from survey respondents. Respondent institution type (i.e., community hospital, academic hospital, pediatric hospital, Veterans Administration / federal hospital, independent reference laboratory, pathology group or clinic) and institution location (i.e., U.S. state, territory, or country) was available for a subset of customers in the CRM for summary analysis. A geographic map of institutions in each U.S. state was created in using a template from TheSlideQuest (https://theslidequest.com/) in PowerPoint (Microsoft 365; Redmond, WA).

Data was tabulated within Qualtrics and further analyzed and displayed using Excel (Microsoft 365; Redmond, WA). Thematic analysis of free text comments was also conducted and included both semantic themes (e.g., the precise wording of a comment) and latent themes (e.g., an underlying concept in a comment) [20]. Respondent text was assigned to thematic categories and tabulated in Excel, as previously described [21]. Respondent comments displayed in the manuscript were edited for spelling and minor grammatical corrections only.

## Results

Of survey invitations distributed by email to 15,513 individuals, 532 questionnaires (3.4% response rate) were either fully or partially completed. Information on respondent institution’s state, territory, or country was known for 429 respondents (80.6%), with 421 institutions in this subset (98.1%) located in the U.S. across 45 states. Eight additional respondents were affiliated with institutions outside the U.S. Distribution of respondent institution location by U.S. state is shown in Figure 1.

**Fig. 1.**
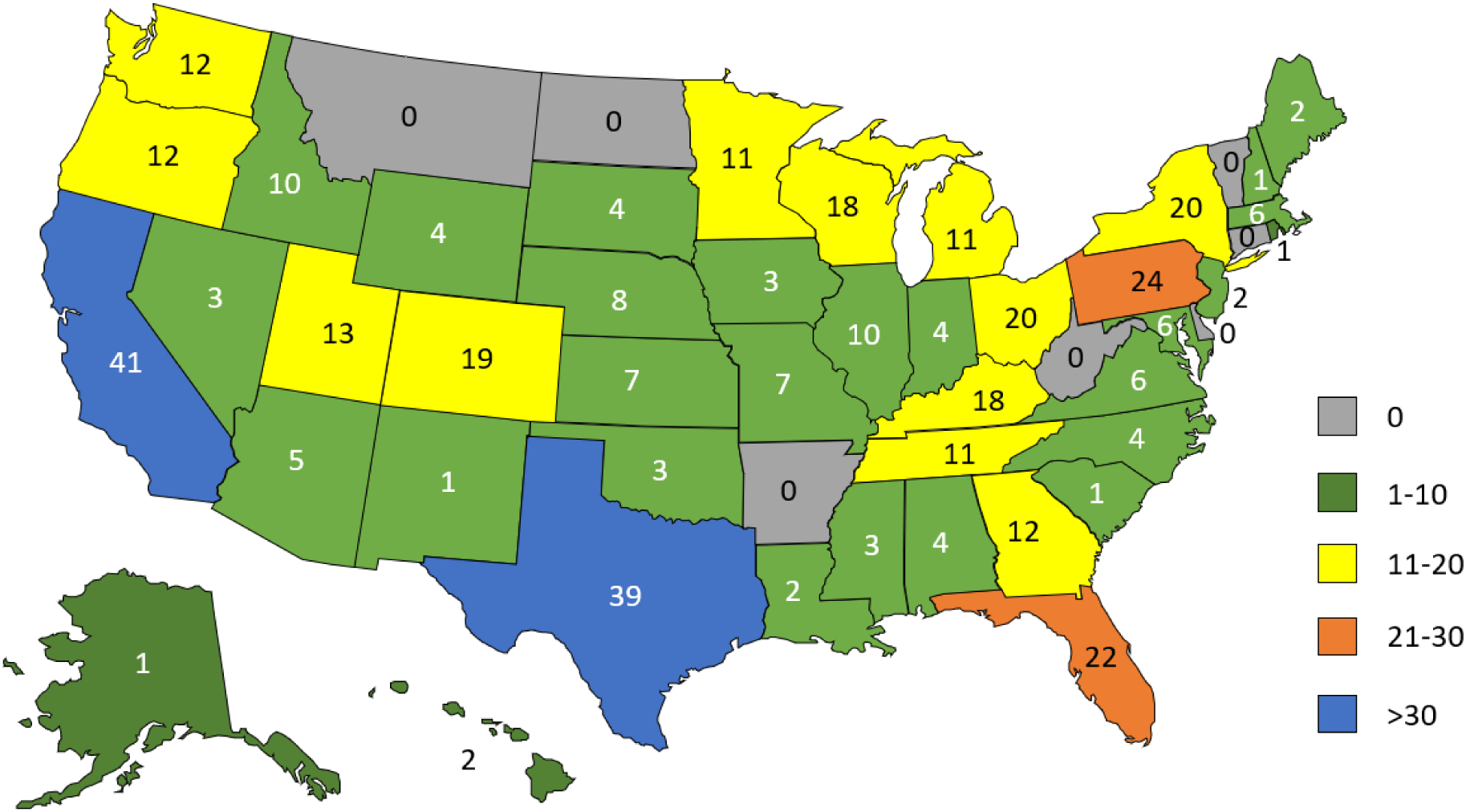
Respondent institution by U.S. state. Map showing respondent institution by state (where available).

Information on respondent institution type was available for 327 respondents (61.5%). Distribution of these institutions by category is shown in Table 1.

**Table 1.** Respondent institution by category.

| **Institution Category** | **Number** | **Percent** |
| --- | --- | --- |
| Hospital (community) | 179 | 54.7% |
| Hospital (academic) | 64 | 19.6% |
| Independent reference laboratory | 40 | 12.2% |
| Hospital (children's) | 25 | 7.6% |
| Hospital (Veterans Administration/federal) | 10 | 3.1% |
| Pathology group or clinic | 9 | 2.8% |

Respondents were asked in the questionnaire to specify a job that best describes their roles within their respective organization. A categorized list of respondent job roles is shown in Table 2. The most common job categories selected by respondents included lab manager or supervisor (153, 28.8%), lab director (97, 18.2%), lab employee (med tech, lab tech, lab assistant; 83, 15.6%), and medical director, pathologist, physician, clinician, or PhD scientist (68, 12.8%).

**Table 2.** Job categories of survey respondents.

| **Job Category** | **Number** | **Percent** |
| --- | --- | --- |
| Lab manager or supervisor | 153 | 28.8% |
| Lab director | 97 | 18.2% |
| Lab employee (med tech / lab tech / lab assistant) | 83 | 15.6% |
| Medical director, pathologist, physician, clinician, or PhD scientist | 68 | 12.8% |
| Send-out / referral testing | 29 | 5.5% |
| Quality and compliance | 25 | 4.7% |
| Executive (CEO, CFO, etc.) | 23 | 4.3% |
| IT / LIS | 13 | 2.4% |
| Customer service / support service | 10 | 1.9% |
| Office: executive assistant, administrative assistant, etc. | 4 | 0.8% |
| Specimen processing / receiving | 1 | 0.2% |
| Supply chain / ancillary services | 1 | 0.2% |
| Other | 25 | 4.7% |
| **Total** | **532** |  |

Abbreviations: CEO, chief executive officer; CFO, chief financial officer; IT, information technology; LIS, laboratory information system

503 respondents then answered a question regarding whether they support the FDA’s proposed rule to regulate LDTs as medical devices. A majority (360, 71.6%) responded ‘no,’ 41 (8.2%) responded ‘yes,’ and 102 (20.3%) responded that they either had no opinion (69, 13.7%) or did not know whether they support the proposed rule (33, 6.6%) (Fig 2a).

**Fig. 2.**
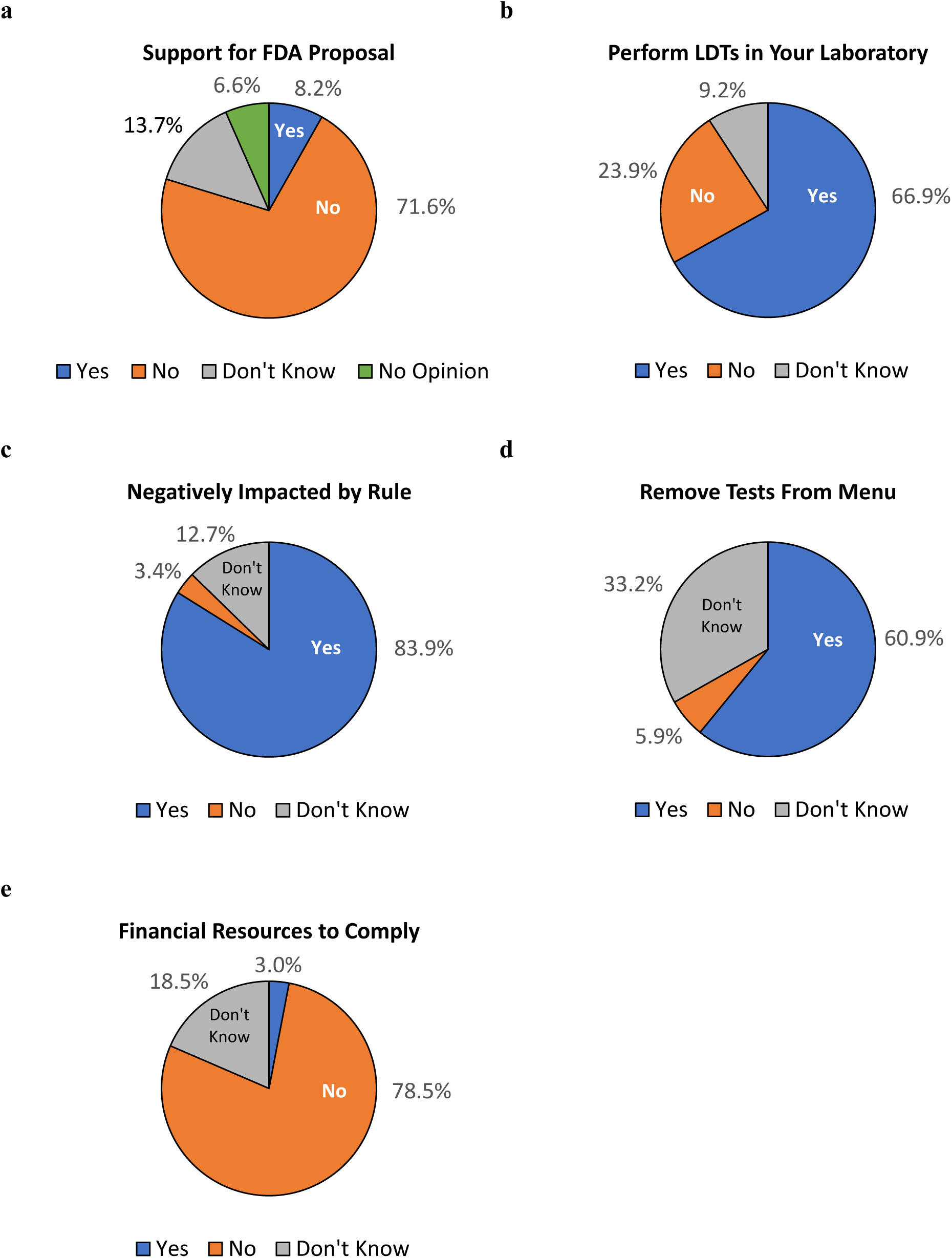
Responses to survey questions. Respondents were asked (a) whether they support the FDA proposal to regulate LDTs, (b) whether they perform LDTs within their laboratories, (c) if their laboratories would be negatively impacted by the proposed rule if enacted, (d) if they anticipate having to remove tests from their laboratory menus if the proposed rule is enacted, and (e) whether they have the financial resources to pay for FDA user fees. Questions for (c), (d), and (e) were asked only of respondents whose laboratories performed LDTs.

489 respondents then answered a question regarding whether their laboratory performs LDTs. 327 (66.9%) responded ‘yes,’ 117 (23.9%) responded ‘no,’ and 45 (9.2%) responded ‘don’t know’ (Fig 2b). As the survey was intended to assess how laboratories that perform LDTs would respond to the proposed FDA rule if enacted, a response of ‘no’ or ‘don’t know’ ended the survey for those respective respondents.

Respondents whose laboratories perform LDTs were then asked whether the FDA’s proposed rule would negatively impact their laboratories (n=322 responses). 270 (83.9%) responded ‘yes,’ 11 (3.4%) responded ‘no,’ and 41 (12.7%) responded ‘don’t know’ (Fig 2c). Respondents were also asked whether they anticipate having to remove tests from their menu if the proposed rule is enacted (n=304 responses). A majority, 185 (60.9%) responded ‘yes,’ 101 (33.2%) responded ‘don’t know,’ and 18 (5.9%) responded ‘no’ (Fig 2d). Respondents were also asked whether their laboratories have the financial resources to pay for FDA user fees, with fiscal year 2024 medical device user fees displayed in the question stem – $21,760 per “moderate risk” 510(k) submission and $483,560 per “high risk” premarket authorization submission [22] (n=303 responses). 238 (78.5%) responded ‘no,’ 56 (18.5%) responded ‘don’t know,’ and 9 (3.0%) responded ‘yes’ (Fig 2e).

High levels of concern regarding the potential impact of the proposed rule were expressed by respondents in this survey (Fig 3), with answers of ‘extremely concerned’ or ‘very concerned’ provided across all topics queried, including patient access to essential testing (269 of 305, 88.2%), availability of financial resources to comply with the proposed regulations (264 of 305, 86.6%), impact on innovation (261 of 306, 85.3%), future laboratory send-out costs (252 of 304, 82.9%), increase in test prices (248 of 303, 81.8%), availability of personnel resources to comply with the proposed regulations (247 of 305, 81.0%), and FDA’s ability to implement the proposed regulations (245 of 305, 80.3%). Only a small percentage of respondents (≤6%) chose either ‘not concerned at all’ or ‘slightly concerned’ for any of the topics queried.

**Fig. 3.**
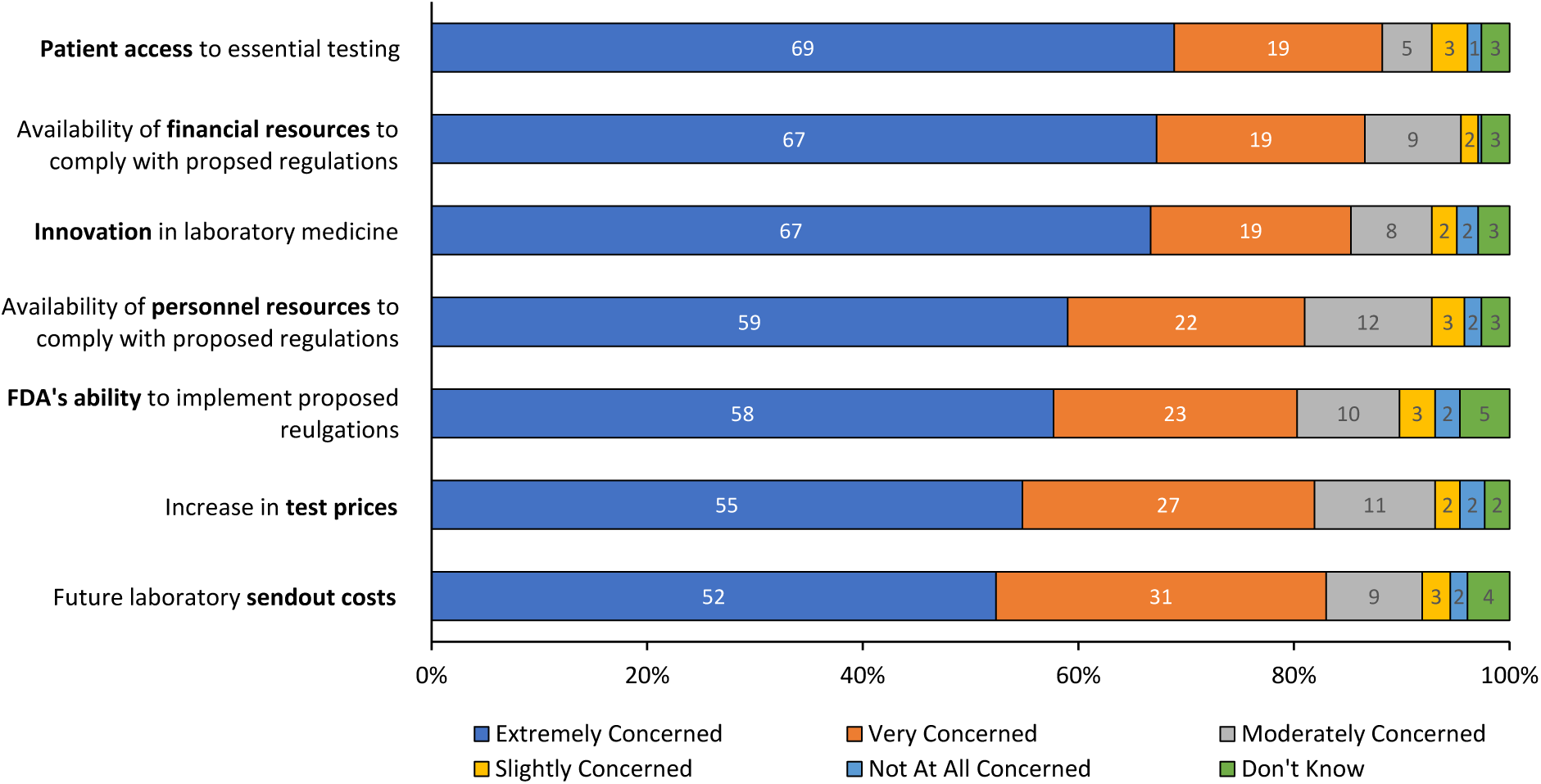
Ratings for level of concern. Stacked bar chart showing respondent level of concern about seven topic areas if the FDA’s proposed rule is adopted. Rating scale: extremely concerned, very concerned, moderately concerned, slightly concerned, not at all concerned, don’t know. Percentages rounded to zero decimal places.

When asked how their laboratories would likely respond to the new regulatory requirements if the FDA adopts the proposed rule, respondents reported the following distribution of strategies regarding potential FDA submissions (n=303 responses): discontinue all existing LDTs that require submissions (48, 15.8%); or alternatively pursue FDA submissions for ‘only a few’ of existing LDTs (50, 16.5%), ‘less than half’ of existing LDTs (29, 9.6%), ‘more than half’ of existing LDTs (41, 13.5%), or ‘all’ existing LDTs (34, 11.2%) (Fig 4). One third of respondents (101, 33.3%) chose ‘don’t know’ to this question, demonstrating that further clarification on the proposed regulations and associated costs will be required to formulate a laboratory’s strategy.

**Fig. 4.**
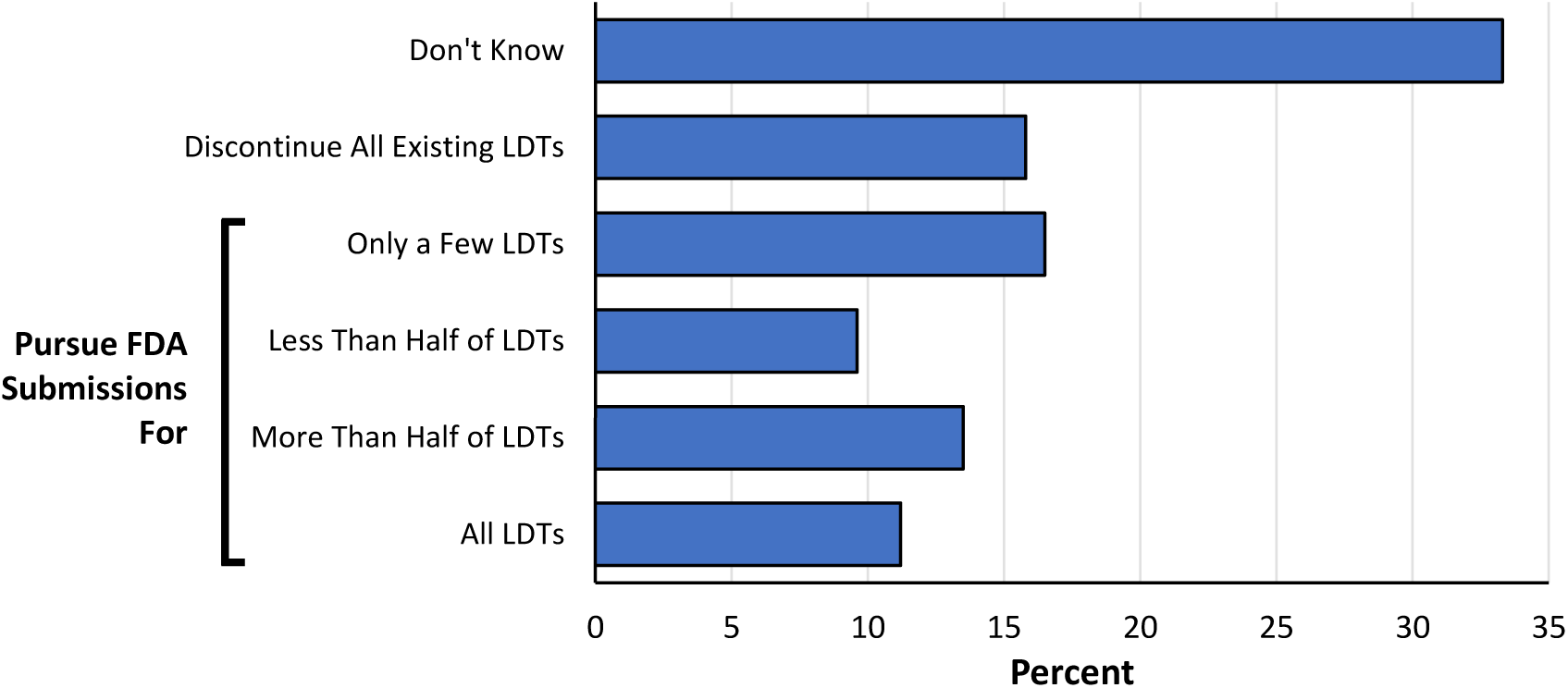
Anticipated response to the proposed rule. Respondents were asked how they think their laboratories would respond to the new regulatory requirements regarding their existing LDTs if the FDA adopts the proposed rule.

Respondents were then asked a question regarding expectations of their reference laboratories if the FDA adopts the proposed rule (n=295 responses; Fig 5). Respondents were asked to select their top two types of support. An equal majority of respondents expected their reference laboratories to advocate on behalf of laboratories to change the FDA rule (175, 59.3%) and to offer testing options for all LDTs that their laboratories discontinue (175, 59.3%), whereas 92 respondents (31.2%) would like consulting services on how to pursue FDA clearance/approval, and 57 respondents (19.3%) would like their reference laboratory to serve as a resource for education about the FDA rule and its implementation. 12 respondents (4.1%) selected ‘other’ and included a variety of additional ideas including participation in litigation against a final rule, manufacturing and distribution of kits, partnership for specialty testing, sharing of protocols and residual specimens for LDT validation purposes, and physician education to alter LDT ordering practices.

**Fig. 5.**
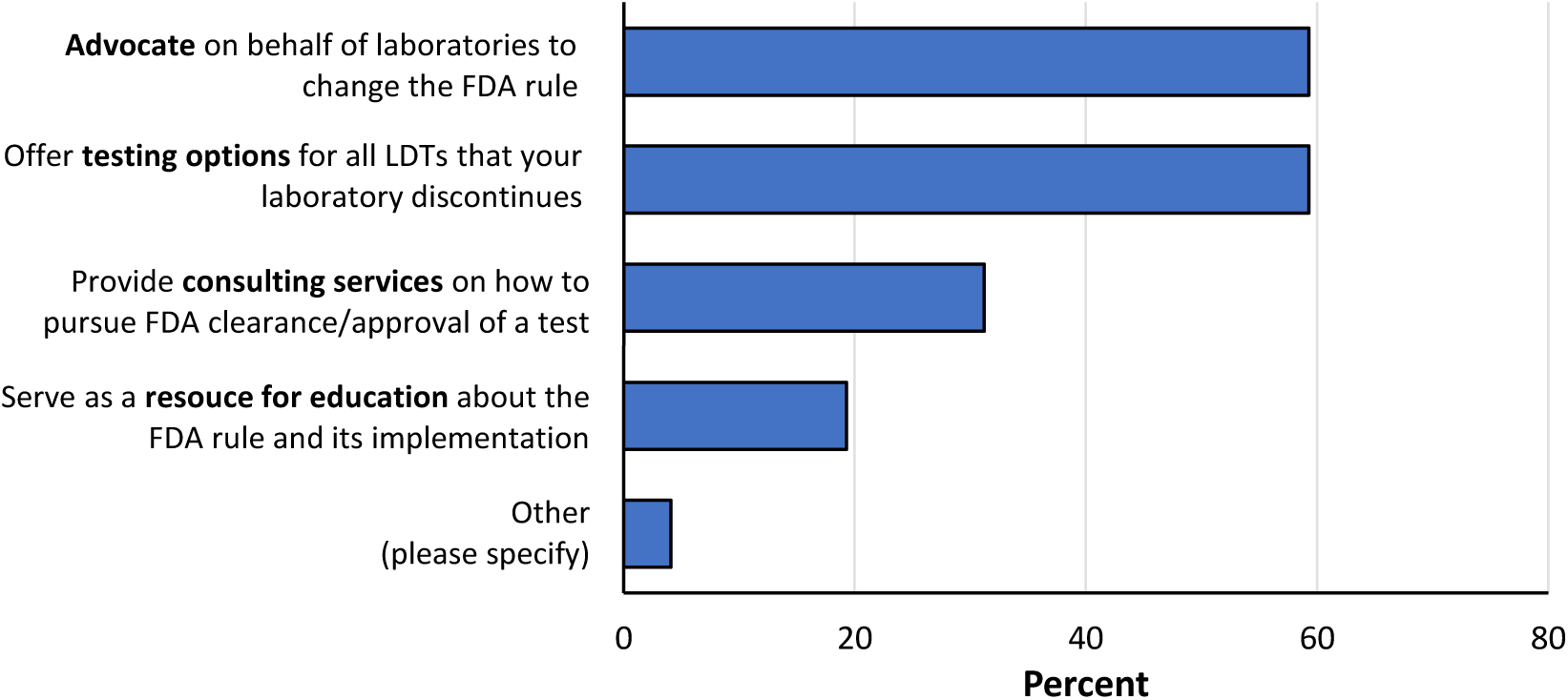
Expected support from reference laboratories. Respondents were asked which types of support they would need from their reference laboratories if the FDA were to adopt the proposed rule. Respondents were instructed to please select their top two types of support.

Respondents were then provided an open text comment field to solicit additional feedback. Additional comments were received from 63 respondents. Of these 63 respondents, 1 (1.6%) was in support of the FDA’s proposed rule, 49 (77.7%) expressed opposition to the proposed rule, and 13 (20.6%) provided feedback that did not address either support or opposition. Multi-part comments were further subdivided by content to reveal a total of 123 distinct comment topics from respondents for thematic analysis. These were then categorized by semantic and latent thematic categories (Table 3). Representative examples of comments are shown in Table 4.

**Table 3.** Thematic analysis of open comments. Thematic categories displayed in the table were mentioned by a minimum of 3 respondents.

| Theme | Number | Percent |
| --- | --- | --- |
| Negative impact to patients |  |  |
| - General | 18 | 14.6 |
| - Pediatric | 4 | 3.3 |
| Cost & negative economic impact | 17 | 13.8 |
| Benefits of alternative regulatory structures (e.g., CLIA, CMS, NYSDOH) | 12 | 9.8 |
| FDA and government overreach | 9 | 7.3 |
| FDA-specific concerns | 8 | 6.5 |
| Discontinuation of testing & laboratory closures | 6 | 4.9 |
| Discipline-specific comments | 5 | 4.1 |
| Gratitude for survey opportunity | 5 | 4.1 |
| LDTs are not medical devices / labs are not manufacturers | 5 | 4.1 |
| Benefits of additional LDT oversight | 4 | 3.3 |
| Supplier challenges with proposed rule | 3 | 2.4 |
| Industry disruption | 3 | 2.4 |
| Other | 24 | 19.5 |

Abbreviations: CLIA, Clinical Laboratory Improvement Amendments; LDT, laboratory-developed test; NYSDOH, New York State Department of Health; TAT, turn-around-time.

**Table 4.**
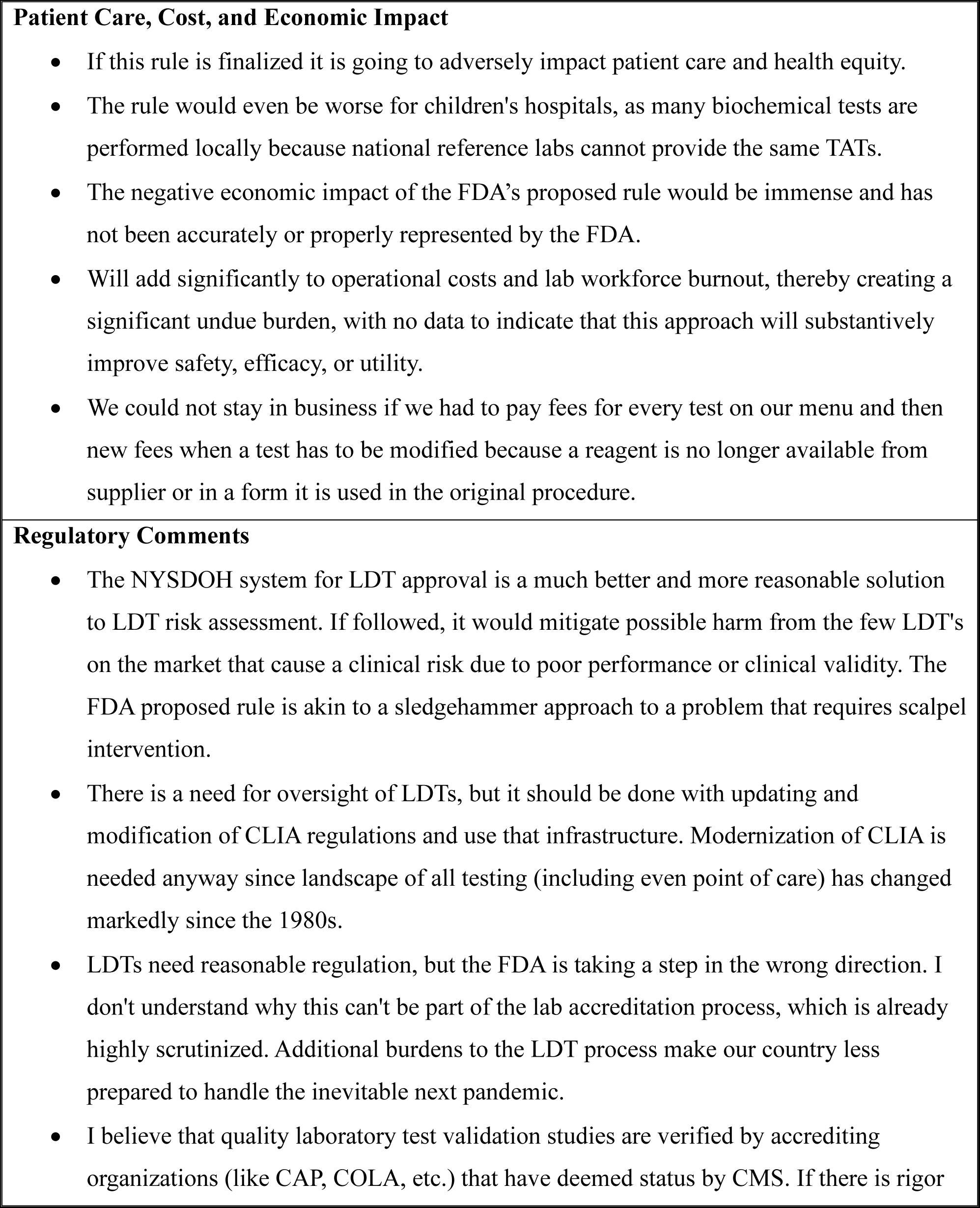
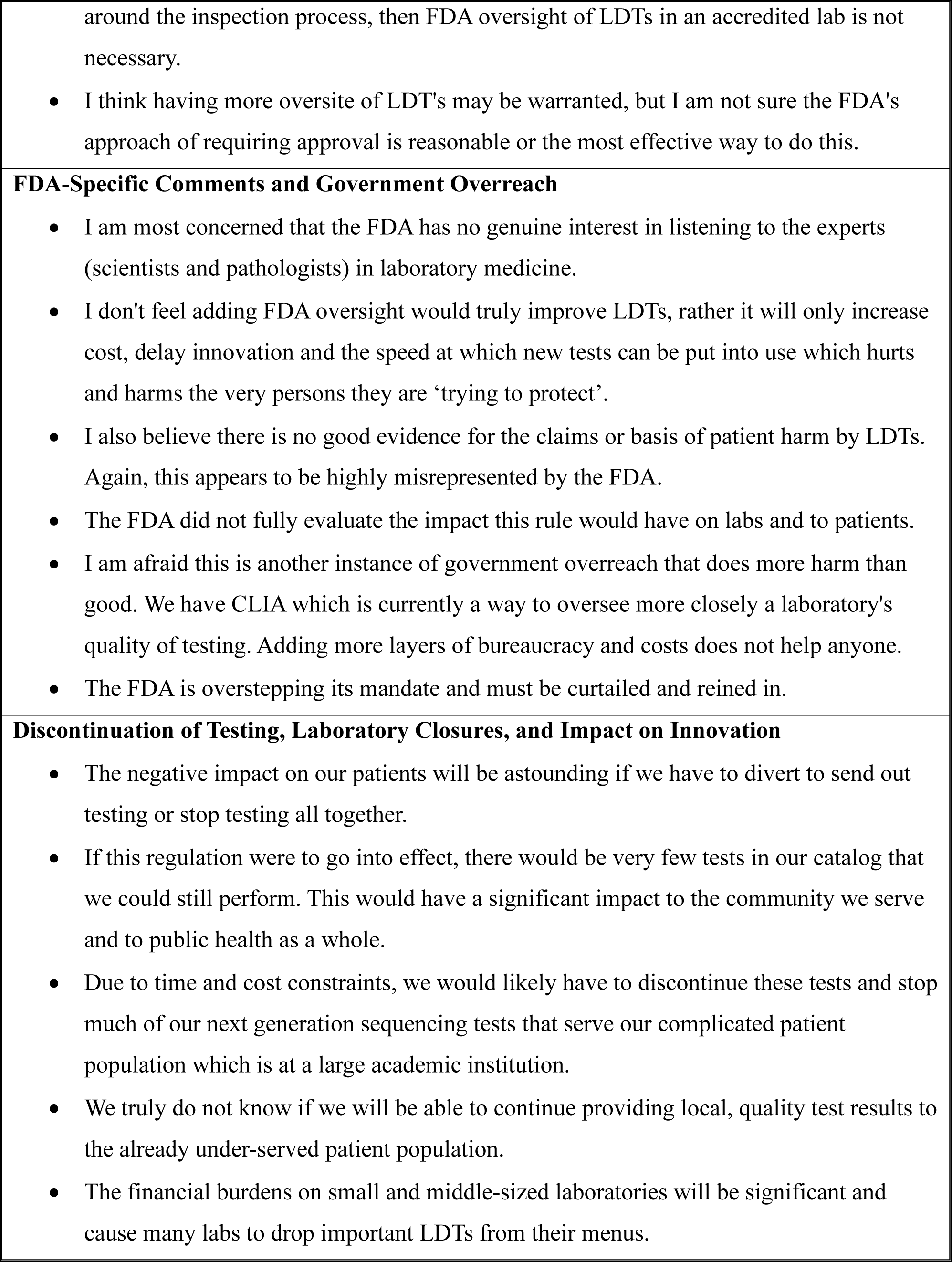
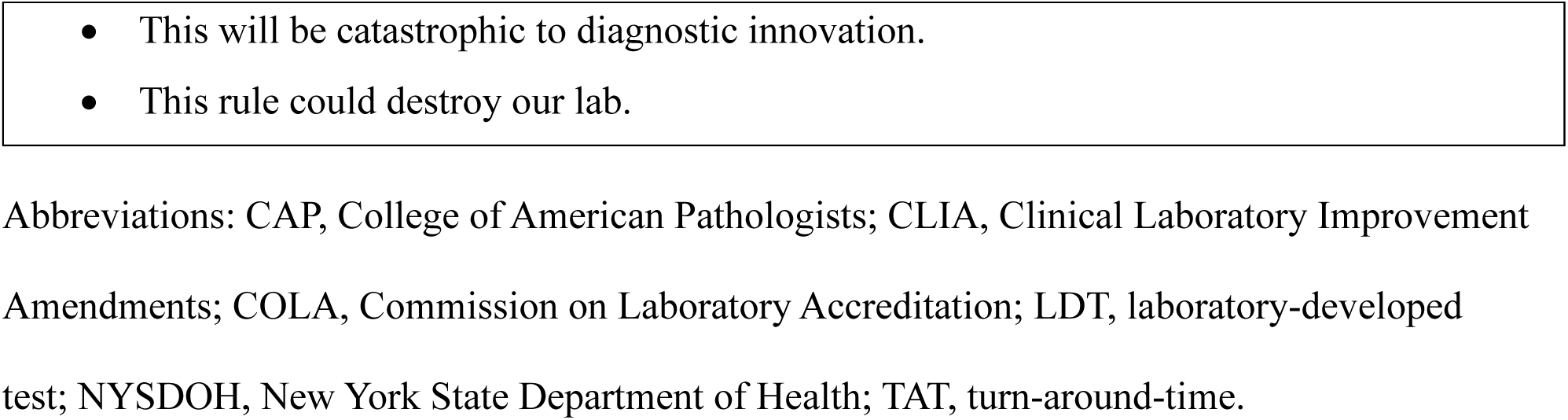
Example Comments from Respondents.

## Discussion

The results of this survey demonstrate that the majority of clinical laboratory respondents are opposed to the FDA’s proposed rule on LDTs and believe that their laboratories and patient care will be negatively impacted if the proposed rule is finalized. Most respondents also reported that they do not have the financial resources to pay for FDA user fees, and that they anticipate having to remove existing clinical tests from their laboratory menus if the proposed rule is enacted. Only a minority of respondents reported that they would likely either submit all of their LDTs, or more than half of their LDTs, for review if the FDA adopts the proposed rule. The majority of respondents also expressed strong concern regarding the proposed rule, including its impact on patient access to essential testing, financial and personal resources necessary to comply with the proposed rule, innovation, the FDA’s ability to implement the proposed rule, and anticipated increases in send-out costs and test prices.

Statutory language in the Medical Device Amendments of 1976 (related to records and reports on devices) stipulates that the agency “shall not impose requirements unduly burdensome to a device manufacturer, importer, or distributor taking into account his cost of complying with such requirements and the need for the protection of the public health and the implementation of this Act” [23]. As such, the FDA has a legal obligation to consider the cost of proposed regulations in the context of this “unduly burdensome” standard. While the FDA released a regulatory impact analysis in October 2023 to review the purported costs and benefits to society with the proposed rule [24], many organizations (including our own) have noted in public comments that the FDA’s analysis includes material errors and assumptions that dramatically under-estimate cost and over-estimate benefit of the proposed rule to society [25, 26]. Negative impact to clinical laboratories and patient care was also not thoroughly assessed in the regulatory impact analysis. The present survey provides additional information regarding the perceived burden of the proposed rule on the clinical laboratory community that should be considered by the FDA, the Department of Health and Human Services (HHS), and the White House Office of Information and Regulatory Affairs (OIRA) prior to advancing any final rule, given the potential of the proposed rule for unanticipated negative impacts to clinical laboratory testing and public health [27, 28].

Survey results demonstrated that 66.9% of respondents perform LDTs within their laboratories. The limited public information on the number and extent of LDTs in use within clinical laboratories has been one justification proposed by the FDA and other groups for additional oversight [24, 29]. This does not mean that such information does not exist, however, but rather it is not being collected by federal agencies in a manner that would better inform the public [30]. For example, and as previously discussed [30], all clinical laboratories accredited by the College of American Pathologists (a CLIA-approved accreditation organization) are required to maintain a list of LDTs currently in use [31], although the Centers for Medicare and Medicaid Services (CMS) does not collect this information from the CAP. Additionally, all laboratories accredited by the New York State Department of Health (NYSDOH) are subject to an LDT review and approval process prior to use [32]. NYSDOH also maintains a searchable public database of all approved LDTs offered by its accredited laboratories [33]. Lastly, CMS requires information on assay manufacturers in clinical laboratory CLIA applications [34]. Ultimately, there are already several non-FDA mechanisms for gathering information on how many LDTs are currently in use in the U.S., although CMS has not been active in either collecting or analyzing this information to assist in the current public discourse.

The question of ‘how many’ different types of LDTs are available across clinical laboratories is often confused with the concept of ‘how frequently’ they are ordered by clinicians. In a retrospective study of real-world clinician test orders at a large academic health system, we previously demonstrated that LDTs are ordered far less frequently than is typically portrayed in public discussion [30]. For example, over one year of all test orders in our health system, approximately 93.9% of clinician laboratory orders were for FDA-cleared/approved assays, while only 3.9% of clinician orders were for LDTs, and 2.3% of clinician orders were for standard methods (e.g., differential cell counts, erythrocyte sedimentation rate, urinalysis, Gram stains, etc.) [30]. Many LDTs were associated with test modifications, including alternative specimen types for which the original assay manufacturer did not seek FDA-clearance/approval in its submissions. Under the FDA’s proposed rule, the burden for regulatory submissions with common test modifications would fall upon clinical laboratories.

IVD manufacturers typically only distribute FDA-cleared/approved assays that are commercially viable, based on expected order volumes for relatively common disorders and/or use cases. LDTs, however, are often used in situations where FDA-cleared/approved alternatives do not exist, or alternatively when LDTs provide superior analytical performance for a desired use as noted by clinical guidelines. These LDT use cases often represent relatively low-test order volumes in hospital settings, making potential compliance costs with additional regulatory oversight and FDA user fees particularly burdensome. The present survey, for example, demonstrates that only 3.0% of clinical laboratory respondents believe that they have the financial resources to comply with existing FDA user fees, and many, if not most, would not qualify for small business discounts as they are incorporated under the hierarchy of much larger health systems. A regulatory structure that is cost-prohibitive will unfortunately result in discontinuation of testing options in many settings.

Survey respondents also provided valuable perspectives on what they expect from their reference laboratory partners if the proposed rule is enacted. For example, many respondents (59.3%) indicated that they will expect their reference laboratory to offer testing options for all LDTs that their laboratories discontinue. While reference laboratory testing provides an essential service in supporting diagnostic testing needs of providers and patients, transportation of specimens to external laboratories – for testing that could otherwise be routinely performed in-house and closer to the patient – introduces previously avoidable delays in diagnosis and treatment decisions. The negative impact of testing delays to patient care was not, however, factored into the FDA’s regulatory impact analysis and deserves careful consideration prior to enactment of any proposed rule.

Many respondents also indicated that they would expect their reference laboratory partner to advocate for changes to the FDA rule on behalf of the respondent laboratory if the proposed rule is adopted. Additional respondents emphasized the desire for education and consultation regarding FDA submissions. Education and advocacy are important elements in reference laboratory partnerships, and this can include a spectrum of activities, including publications [8, 35–38], podcasts [39], webinars and online resources [40–42], and public comments and advocacy letters [25, 38, 43]. An informed clinical laboratory community can better advocate for the needs of its patients, and the survey demonstrates that reference laboratories play an important and ongoing role in these efforts.

Thematic analysis of clinical laboratory respondent free-text responses also revealed significant concerns regarding the proposed rule’s impact across multiple domains, including a negative impact to patient care, costs and economic impacts, the benefits of alternative regulatory structures, anticipated discontinuation of testing and laboratory closures, as well as concerns specific to the FDA and an overall perception of government overreach. Numerous comments were directly critical of the FDA and its approach to LDTs, highlighting a considerable lack of trust in the agency by some respondents, at least in the context of the current proposed rule. Potential factors that may contribute to these concerns include clinical laboratorian interactions with the FDA during the COVID-19 pandemic [44, 45], unresolved legal questions regarding jurisdiction over LDTs [8], FDA public statements critical of LDTs (and therefore overall quality of testing performed in respondent laboratories) [46], and a disconnect between FDA cost estimates and the financial challenges and staffing constraints within which most clinical laboratories and hospitals operate [47, 48]. Deriving a better understanding of the causes for these concerns will be critically important for the success of any future regulatory proposals and should therefore be the subject of further study by the FDA and HHS.

There are several limitations to the present study. The survey distribution included only individuals within one large national reference laboratory’s customer contact list. The results represent the opinions of respondents who agreed to participate in this survey, and there is the potential for selection bias in responses (e.g., individuals who have a vested interest in LDTs may have been more likely to complete the survey). As results were anonymized, we could not categorize the job classifications of those who responded ‘other’, nor did we exclude any job classifications to create a more comprehensive overview of clinical laboratory responses. Finally, as multiple extension requests during the public comment period were denied by the FDA, the present results could not be submitted in the public comment submission system, as the portal was closed at the time of study completion. We therefore request that the FDA continue to solicit feedback and work with stakeholders to better understand the negative implications of the proposed rule on clinical laboratories and the public health.

## Disclosures

All authors have completed the ICMJE uniform disclosure form at www.icmje.org/coi_disclosure.pdf and declare: no support from any organization for the submitted work; JRG serves as chief medical officer at ARUP Laboratories and is an employee of the University of Utah; LS is an employee of ARUP Laboratories; LAC is an employee of ARUP Laboratories; no other relationships or activities that could appear to have influenced the submitted work. This study did not receive any external funding.

## Data Availability

The data that support the findings of this study are available from the corresponding author, JRG, upon reasonable request.

## Survey Questionnaire

**Which of the following job categories best describes your role within the organization?**

- Customer Service / Support Service
- Executive (CEO, CFO, etc.)
- IT/LIS
- Lab Director
- Lab Employee (Med Tech / Lab Tech / Lab Assistant)
- Lab Manager or Supervisor
- Medical Director, Pathologist, Physician, Clinician, or PhD Scientist
- Office: Executive Assistant, Administrative Assistant, etc.
- Quality and Compliance
- Sendout / Referral Testing
- Specimen Processing / Receiving
- Supply Chain / Ancillary Services
- Other (*please specify*)

**Do you support the FDA’s proposed rule to regulate laboratory-developed tests (LDTs) as medical devices?**

- Yes
- No
- I don’t have an opinion about this proposed rule
- Don’t know

**Does your laboratory perform laboratory-developed tests (LDTs)?**

- Yes
- No
- Don’t know

**Do you believe your laboratory will be negatively impacted by the FDA’s proposed rule?**

- Yes
- No
- Don’t know

**Please rate your level of concern about the following if the FDA’s proposed rule is adopted:** *[Likert Scales; Not at all concerned, Slightly concerned, Moderately concerned, Very concerned, Extremely concerned, Don’t know]*

- Patient access to essential testing
- Availability of financial resources to comply with proposed regulations
- Availability of personnel resources to comply with proposed regulations
- Innovation in laboratory medicine
- FDA’s ability to implement proposed regulations
- Future laboratory send-out test costs
- Increase in test prices
- Other (*please specify*)

**Do you anticipate having to remove tests from your menu if the proposed rule is enacted?**

- Yes
- No
- Don’t know

**If the FDA were to adopt the proposed rule, how do you think your laboratory would likely respond to the new regulatory requirements?**

- We would pursue FDA submissions for **all** of our existing LDTs
- We would pursue FDA submissions for **more than half** of our existing LDTs
- We would pursue FDA submissions for **less than half** of our existing LDTs
- We would pursue FDA submissions for **only a few** of our existing LDTs
- We would **discontinue all** of our existing LDTs that require FDA submissions
- Don’t know

**In your opinion, does your laboratory have the financial resources to pay for FDA user fees? For example, current medical device user fees are $21,760 per “moderate risk” 510(k) submission and $483,560 per “high risk” premarket authorization submission.**

- Yes
- No
- Don’t know

**Which types of support would you need from your reference laboratories if the FDA were to adopt the proposed rule? (*Please select the top 2 types of support*)**

- Offer testing options for all LDTs that your laboratory discontinues
- Serve as a resource for education about the FDA rule and its implementation
- Provide consulting services on how to pursue FDA clearance/approval of a test
- Advocate on behalf of laboratories to change the FDA rule
- Other (*please specify*)

**Do you have any additional feedback that you did not have an opportunity to share in the survey?** [*open text field*]

## Notes

### Funding Statement

This study did not receive any funding

### Author Declarations

IRB of University of Utah gave a determination of exemption for this work

### Summary of Updates

Spelling correction in Figure 2.

